# Percutaneous coronary intervention with drug-eluting stent is associated with better survival than coronary artery bypass grafting in Taiwan dialysis patients

**DOI:** 10.1101/2023.05.17.23290146

**Authors:** Szu-Yu Pan, Ju-Yeh Yang, Nai-Chi Teng, Yun-Yi Chen, Shi-Heng Wang, Chien-Lin Lee, Kang-Lung Chen, Yen-Ling Chiu, Shih-Ping Hsu, Yu-Sen Peng, Yung-Ming Chen, Shuei-Liong Lin, Likwang Chen

## Abstract

**Rationale and Objective:** To study the comparative effectiveness of percutaneous coronary intervention with drug-eluting stent and coronary artery bypass grafting in dialysis patients.

**Study Design:** Retrospective observational cohort study.

**Setting and Participants:** This population-based study identified dialysis patients hospitalized for coronary revascularization between January 1, 2009 and December 31, 2015 in Taiwan National Health Insurance Research Database.

**Exposures:** Percutaneous coronary intervention with drug-eluting stent versus coronary artery bypass grafting.

**Outcomes:** All-cause mortality, in-hospital mortality, and repeat revascularization.

**Analytical Approach:** Propensity scores were used to match patients. Cox proportional hazards models and logistic regression models were constructed to examine associations between revascularization strategies and mortality. Interval Cox models were fit to estimate time-varying hazards during different periods.

**Results:** A total of 1,840 propensity score-matched dialysis patients were analyzed. Coronary artery bypass grafting was associated with higher in-hospital mortality (coronary artery bypass grafting vs. percutaneous coronary intervention with drug-eluting stent, crude mortality rate 12.5% vs. 3.3%; adjusted odds ratio 5.22; 95% confidence interval [CI] 3.42-7.97; p < 0.001), and longer hospitalization duration (median [interquartile range], 20 [14-30] days vs. 3 [2-8] days, p < 0.001). After discharge, repeat revascularization, acute coronary syndrome, and repeat hospitalization all occurred more frequently in the percutaneous coronary intervention with drug-eluting stent group. Importantly, with a median follow-up of 2.8 years, coronary artery bypass grafting was significantly associated with a higher risk of all-cause overall mortality (adjusted hazard ratio 1.19, 95% CI 1.05-1.35, p < 0.01) in the multivariable Cox proportional hazard model. Sensitivity and subgroup analyses yielded consistent results.

**Limitations:** Observational study with mainly Asian ethnicity.

**Conclusions:** Percutaneous coronary intervention with drug-eluting stent may be associated with better survival than coronary artery bypass grafting in dialysis patients. Future studies are warranted to confirm this finding.

## Introduction

In patients with end-stage kidney disease (ESKD), cardiovascular mortality is the leading cause of death.^1^ Coronary artery disease (CAD) is an important etiology of cardiovascular mortality. Coronary revascularization with either coronary artery bypass grafting (CABG) or percutaneous coronary intervention (PCI) is a critical therapeutic strategy in addition to medical treatment.

In the context of non-emergent multi-vessel CAD among the general population, CABG is associated with better long-term outcome than PCI.^2-5^ However, uremic milieu exposes ESKD patients to several non-traditional cardiovascular risk factors, and the benefit of CABG over PCI is less clear.^6,7^ In patients with chronic kidney disease (CKD), a pooled analysis of patient-level data from RCTs reported that CABG, compared with PCI, did not improve survival, although both subsequent myocardial infarction and revascularization were reduced.^8^ However, none of the included CKD patients was under dialysis. One observational study, performed by Chang TI and colleagues by using data from the United States Renal Data System (USRDS), reported a worse in-hospital survival for ESKD patients receiving CABG compared with those receiving PCI between 1997 and 2009. However, the long-term outcome was better with CABG.^9^ Mainly based on this study, the European Society of Cardiology and the European Association for Cardio-Thoracic Surgery (ESC/EACTS) guidelines suggest a possibly favorable role of CABG over PCI in dialysis patients with CAD.^10,11^

Importantly, studies in dialysis patients from the United States (US) and Taiwan reported that PCI with DES was associated with not only reduced revascularization but also decreased mortality compared with PCI with bare metal stent (BMS),^12,13^ and the 2018 ESC/EACTS guideline suggested a superior role of DES to BMS in CKD patients. It is thus desirable to examine the comparative effectiveness of CABG and PCI with DES in dialysis patients. However, so far there has been no large-scale RCT demonstrating unequivocal advantage of CABG over PCI with DES in ESKD patients. The landmark SYNTAX and EXCEL trials included only 6 and 3 dialysis patients, respectively.^14-16^ One large observational study using the URSRDS database reported epidemiologic data of survival and repeat revascularization after CABG, BMS, and DES, but did not make a comparison between these strategies.^17^ Another large USRDS study compared CABG with PCI, but the stent types of PCI were not specified.^9^ A recent meta-analysis of observational studies and post-hoc analyses of RCTs identified 801 dialysis patients and reported no difference in all-cause mortality between CABG and PCI with DES.^18^ However, the single largest observational study, including 486 propensity score-matched dialysis patients, reported CABG was associated with a lower risk for mortality and revascularization.^19^ Considering the paucity of evidence, study on this issue is urgently needed.

To analyze the outcome of CABG versus PCI with DES in ESKD patients, we used the Taiwan National Health Insurance Research Database (NHIRD).^20^ Taiwan NHI program has coverage of up to 99% for the whole citizens and the database is representative of the national population. We included 4,165 dialysis patients and analyzed 1,840 propensity score-matched subjects for comparison of CABG versus PCI with DES performed between Jan 1, 2009 to Dec 31, 2015. Surprisingly, CABG was associated with increased both in-hospital and long-term mortality, although repeat revascularization and subsequent acute coronary syndrome (ACS) were reduced.

## Methods

### Study population

Our study used the NHIRD through Applied Health Research Data Integration Service from Taiwan’s National Health Insurance Administration (NHIA). This retrospective cohort study was approved by the Institutional Review Board of National Health Research Institutes, Taiwan (EC1060402-E). The individual information in the NHIRD was encrypted, therefore the requirement to obtain informed consent was waived. We used the data between Jan 1, 2008 and Dec 31, 2017 in the database to identify dialysis patients hospitalized for revascularization by CABG or PCI with DES in Taiwan (eFigure 1 in the **Supplement**). The NHIRD has been successfully used to analyze outcomes of specified populations such as ESKD patients.^13,21^ We set the cohort entry date between Jan 1, 2009 to Dec 31, 2015 to ensure an observation period of at least 1 year before cohort entry and 2 years after revascularization. The index hospitalization was defined as the first admission for coronary revascularization by either CABG or PCI with DES in ESKD patients, and the index date was the date of revascularization. To avoid the coding error frequently encountered in database study, we defined the dialysis population and revascularization intervention with procedure codes, which are directly linked to reimbursement in NHI and less prone to miscoding. The use of procedure codes and material codes in Taiwan NHI was briefly introduced in the Online-Only **Supplement**.

### Outcome

The primary outcome of interest was survival after revascularization. Survival was determined from the index date to death or a censoring date. Patients were censored if they received kidney or heart transplantation after the index date, lost to follow-up, or survived through Dec 31, 2017. To estimate the short-term and long-term effects of revascularization on crude all-cause mortality, we calculated in-hospital mortality rate, and cumulative mortality rates at 1, 2, 3, 4, and 5 years. Kaplan-Meier plot was used to visualize the difference in unadjusted survival between groups. The adjusted odds ratio of CABG over PCI with DES for in-hospital mortality was estimated in the logistic regression model. Adjusted hazard ratios (HRs) for 1, 2, 3, 4, 5-year, and overall mortality were estimated in the Cox proportional hazard model.^22^ Adjusted survival curves based on a Cox model using baseline statement were used to demonstrate adjusted survival probabilities after revascularization. As secondary outcomes, we analyzed the frequency of repeat revascularization, ACS after the index hospitalization, and repeat hospitalization. Repeat revascularization procedures included CABG and PCI with or without DES. ACS included ST-elevation myocardial infarction (STEMI), non-ST elevation myocardial infarction (NSTEMI), and unstable angina (UA).

In the Online-Only Supplement, we provide details for the description of the selection of patients into analysis, covariates, propensity score matching, and statistical analyses.

## Results

### The patients, crude mortality rate, and duration of hospitalization

We identified 4,165 ESKD patients receiving revascularization by either CABG or PCI with DES during the index hospitalization (eFigure 1 in the **Supplement**). Among the 4,165 patients, 1,023 received CABG, and 3,142 received PCI with DES. Compared with patients receiving PCI with DES, patients receiving CABG were younger, more likely to be male, less likely to have an intervention on only one vessel, more likely to have comorbid CAD, congestive heart failure (CHF), and dyslipidemia, less likely to have comorbid cancer, and less likely to have UA or NSTEMI during the index hospitalization (**Table 1**). Notably, Shock or respiratory failure within 24 hours before revascularization was more likely to develop in patients receiving CABG than PCI with DES. In addition, patients were more likely to receive CABG than PCI with DES in hospitals with a high volume of CABG or medical centers. To minimize the inequity of baseline characteristics, we performed a 1:1 propensity score matching and identified 920 matched pairs. After matching, all the covariates were well balanced (**Table 1**).

**Table 1.**
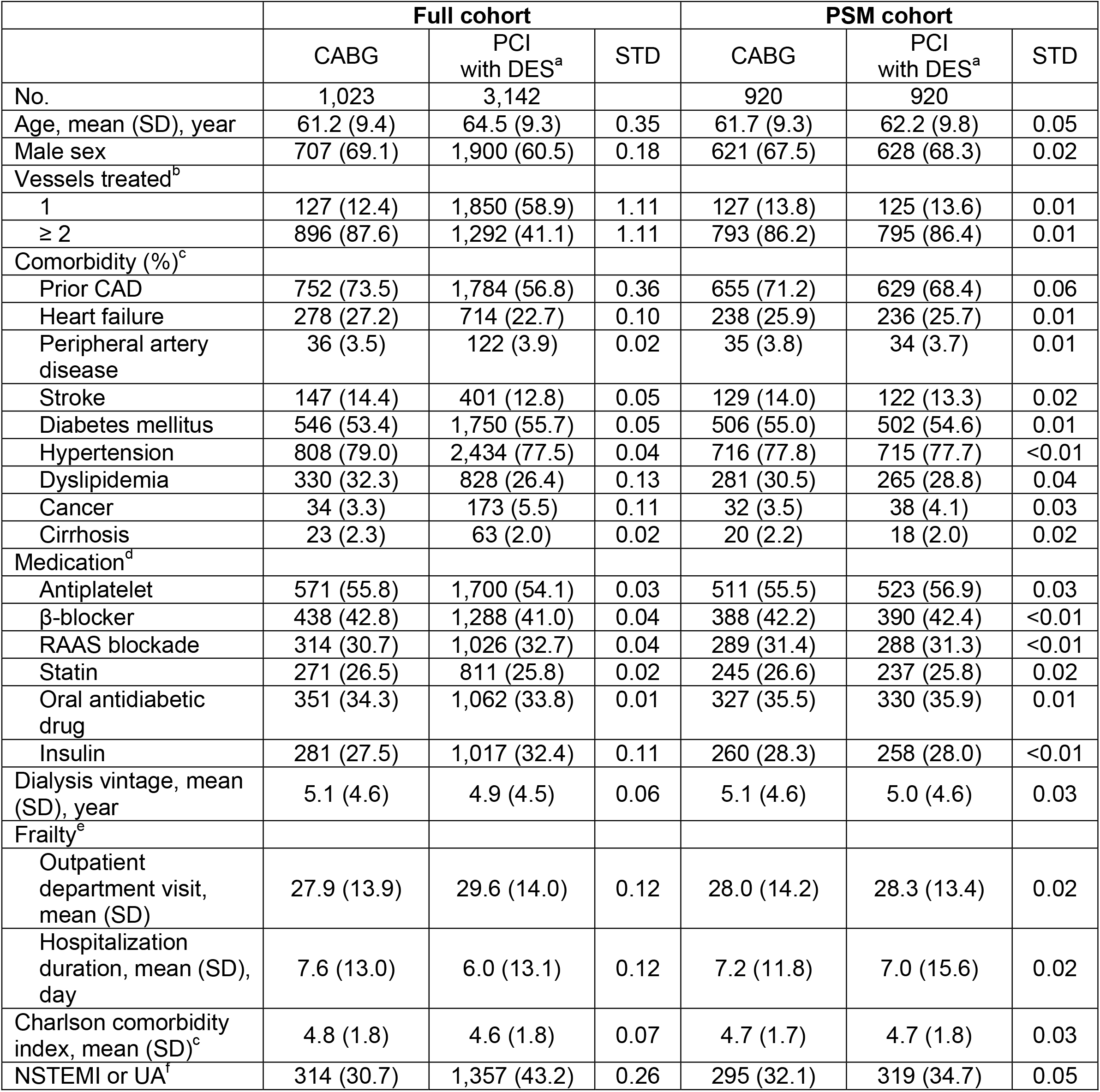
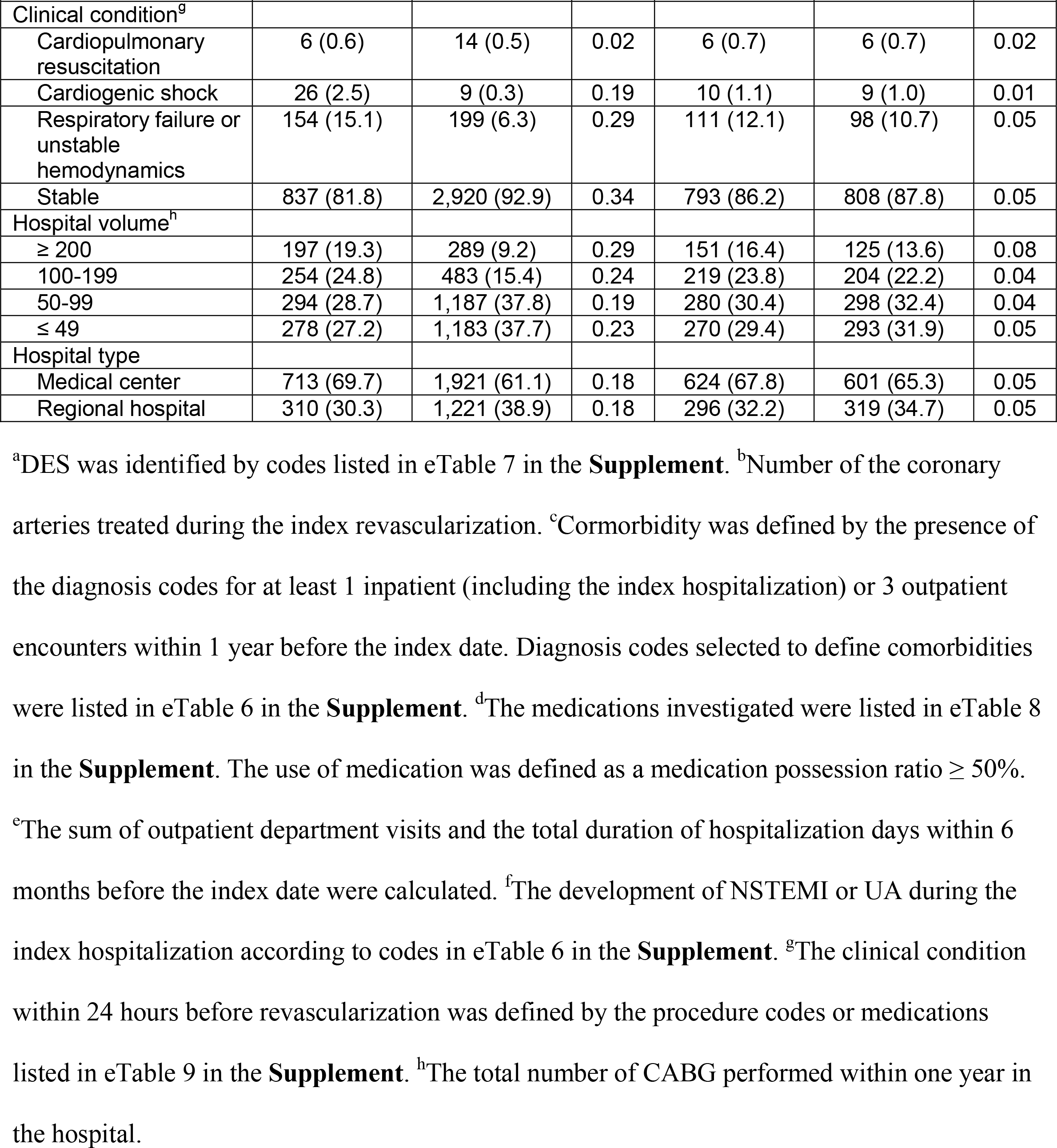
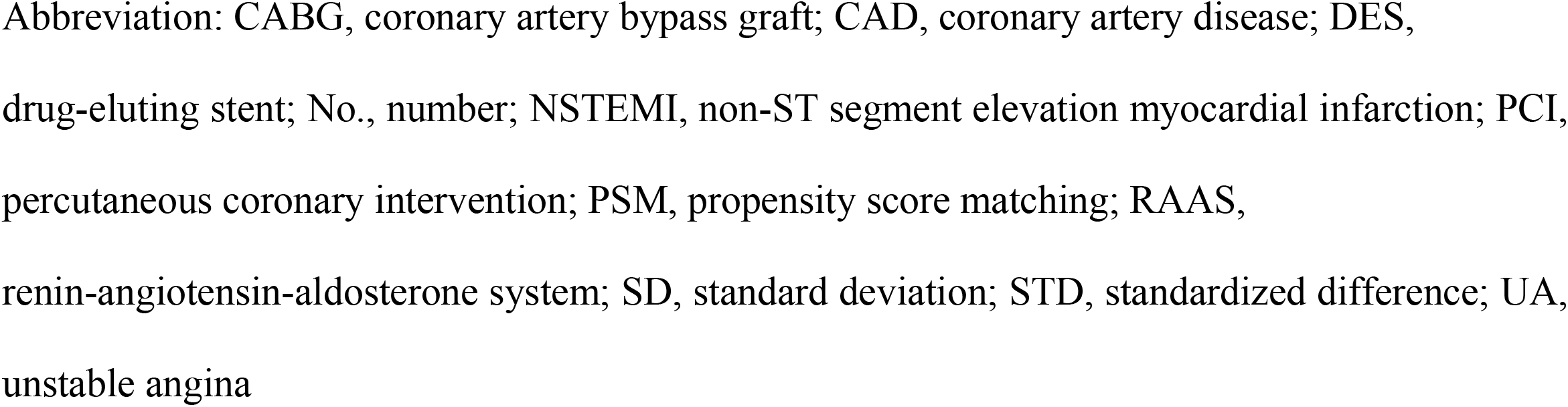
Baseline characteristics of the ESKD patients receiving CABG or PCI with DES in the full and the PSM cohorts.

The median and interquartile range of follow-up duration were 2.7 (0.9-4.4), 2.9 (1.5-4.4), and 2.8 (1.2-4.4) years in the CABG, PCI with DES, and the whole cohort, respectively. The crude mortality rates of matched patients, including mortality rates at the index hospitalization, at 1, 2, 3, 4, and 5 years, were calculated. Surprisingly, the cumulative mortality rates were significantly higher in the CABG than in the PCI with DES group at all the analyzed time points (eTable 1 in the **Supplement**). Kaplan-Meier plot also showed a significant survival advantage in the PCI with DES group (**Figure 1**). The duration of the index hospitalization was longer in the CABG group (CABG 20 [14-30] days vs. PCI with DES 3 [2-8] days, p < 0.001, eTable 1 in the **Supplement**).

**Figure 1.**
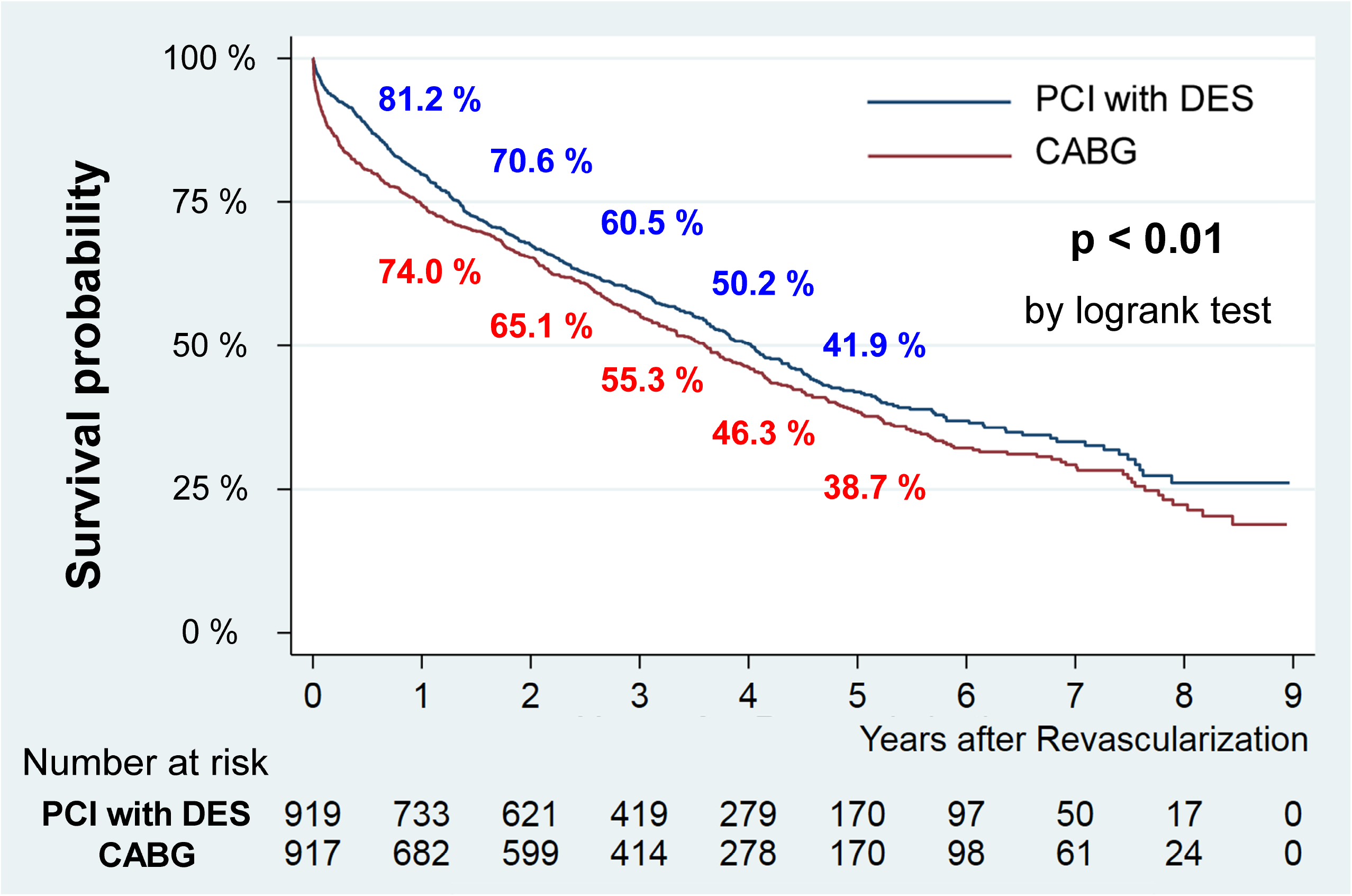
Kaplan-Meier plot for survival in the matched cohort. The Kaplan-Meier plot of survival probabilities in the matched CABG (red line) and PCI with DES (blue line) groups during the study period. In the CABG group, the survival probabilities at 1, 2, 3, 4, and 5 years were 74.0%, 65.1%, 55.3%, 46.3%, and 38.7%, respectively. In the PCI with DES group, the survival probabilities at 1, 2, 3, 4, and 5 years were 81.2%, 70.6%, 60.5%, 50.2%, and 41.9%, respectively. Three patients in the CABG group and one patient in the PCI with DES group died within 1 day after revascularization. p < 0.01 by logrank test. Abbreviation: CABG, coronary artery bypass graft; DES, drug-eluting stent; PCI, percutaneous coronary intervention

### Association between different revascularization strategies and survival

We analyzed in-hospital mortality in the logistic regression model and long-term mortality in the Cox proportional hazard model. Covariates adjusted in the models included age, sex, number of treated coronary vessels, clinical condition before revascularization, UA and NSTEMI during the index hospitalization, utilization of medical resource before the index hospitalization (frailty), comorbidity, medication, hospital type, and hospital volume for CABG. In the adjusted logistic regression model, CABG was strongly associated with increased in-hospital mortality over PCI with DES (adjusted OR 5.22, 95% CI 3.42-7.97, p < 0.001, **Table 2** and eTable 2 in the **Supplement**). Importantly, in the adjusted Cox proportional hazard model during a median follow-up of 2.8 years, CABG was also associated with increased overall all-cause mortality over PCI with DES (adjusted HR 1.19, 95% CI 1.05-1.35, p < 0.01, **Table 2** and eTable 3 in the **Supplement**). The HRs for all-cause mortality at 1, 2, 3, 4, and 5 years were also increased in the CABG group (**Table 2**). The adjusted survival curves for overall survival also showed superior survival in the PCI with DES group (eFigure 2 in the **Supplement**).

**Table 2.** Comparison of mortality risks between the 2 treatment groups.

|  | **OR or HR** | **95% CI** | **p value** |
| --- | --- | --- | --- |
| In-hospital mortality | 5.22 | 3.42-7.97 | <0.001 |
| 1-year mortality | 1.54 | 1.26-1.87 | <0.001 |
| 2-year mortality | 1.31 | 1.11-1.54 | <0.01 |
| 3-year mortality | 1.24 | 1.08-1.44 | <0.01 |
| 4-year mortality | 1.20 | 1.05-1.37 | <0.01 |
| 5-year mortality | 1.18 | 1.04-1.34 | 0.01 |
| Overall mortality | 1.19 | 1.05-1.35 | <0.01 |

The adjusted OR for in-hospital mortality was estimated by the multivariable logistic regression.
The adjusted HRs for 1-, 2-, 3-, 4-, and 5-year mortality rates, and overall mortality were estimated by the multivariable Cox proportional hazard model. PCI with DES was set as the reference group.
Abbreviation: HR, hazard ratio; OR, odds ratio

Notably, although CABG was associated with higher overall mortality, PCI with DES was associated with more subsequent ACS, more repeat revascularization, and more repeat hospitalization (eTable 4 in the **Supplement**).

### Sensitivity and subgroup analyses

With regard to overall mortality (**Table 3**) and in-hospital mortality (eTable 5 in the **Supplement**), similar results were yielded in the sensitivity analyses of the unmatched cohort, after exclusion of mortality within 3 days after revascularization, after exclusion of patients with UA or NSTEMI during the index hospitalization, after exclusion of patients with unstable clinical conditions within 24 hours before revascularization, and after exclusion of patients receiving intervention on only one coronary artery. Because in-hospital mortality negatively impacted long-term survival in patients receiving CABG, we analyzed overall mortality in patients who survived the index hospitalization and set the date of discharge from index hospitalization as day 0 in the Cox model. Surprisingly, CABG was not associated with better long-term survival over PCI with DES even after the exclusion of patients who died during the hospitalization (HR 1.02, 95% CI 0.89-1.17, p = 0.74). Interval Cox model also revealed that PCI with DES was non-inferior to CABG during different time periods, and CABG was associated with higher mortality hazard in the first 2 years after revascularization, likely due to higher perioperatively in-hospital mortality risk (**Table 3**). Notably, when repeat revascularization was included in the composite outcomes, CABG was significantly associated with decreased HR risks compared with PCI with DES (**Table 3**).

**Table 3.**
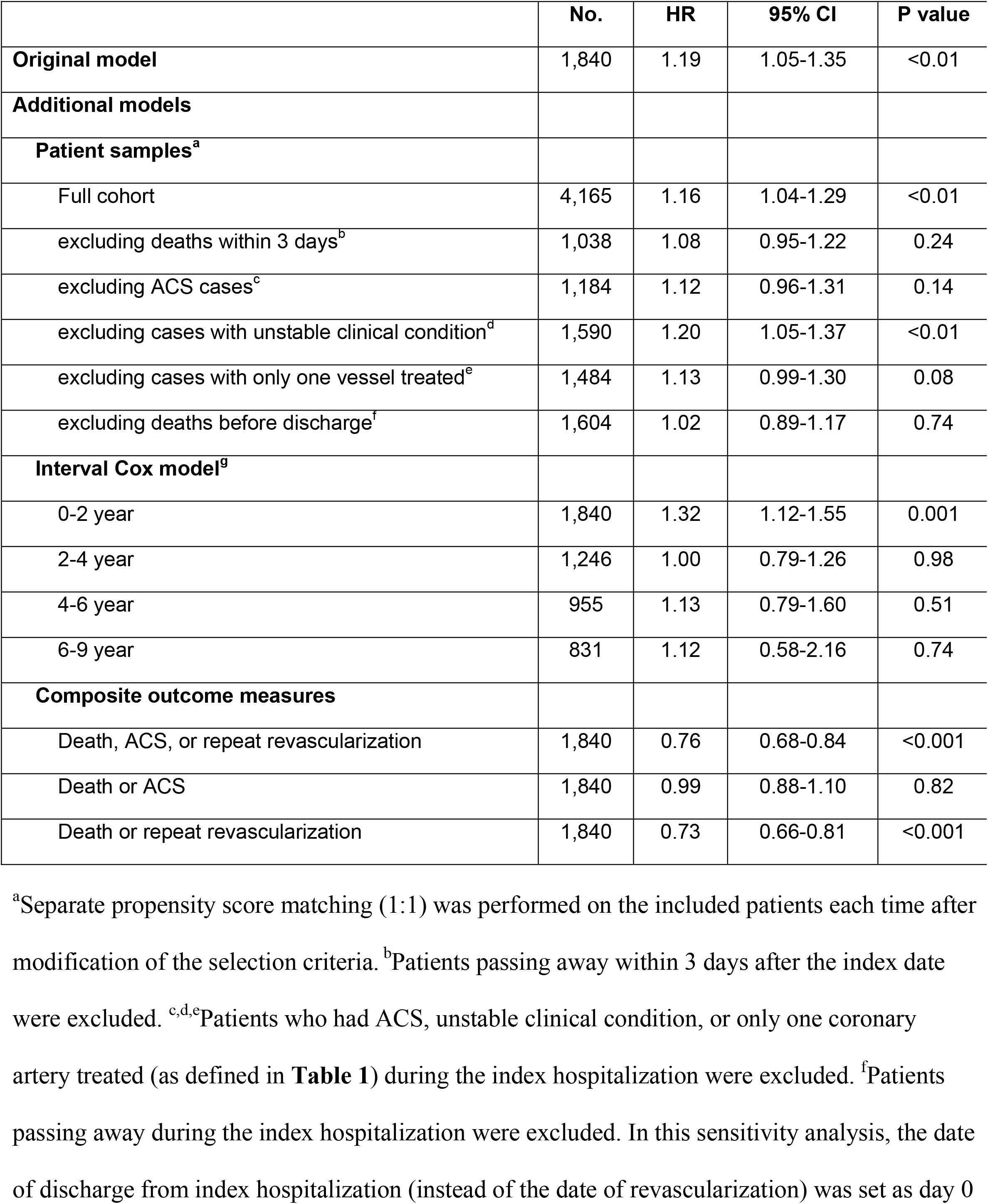
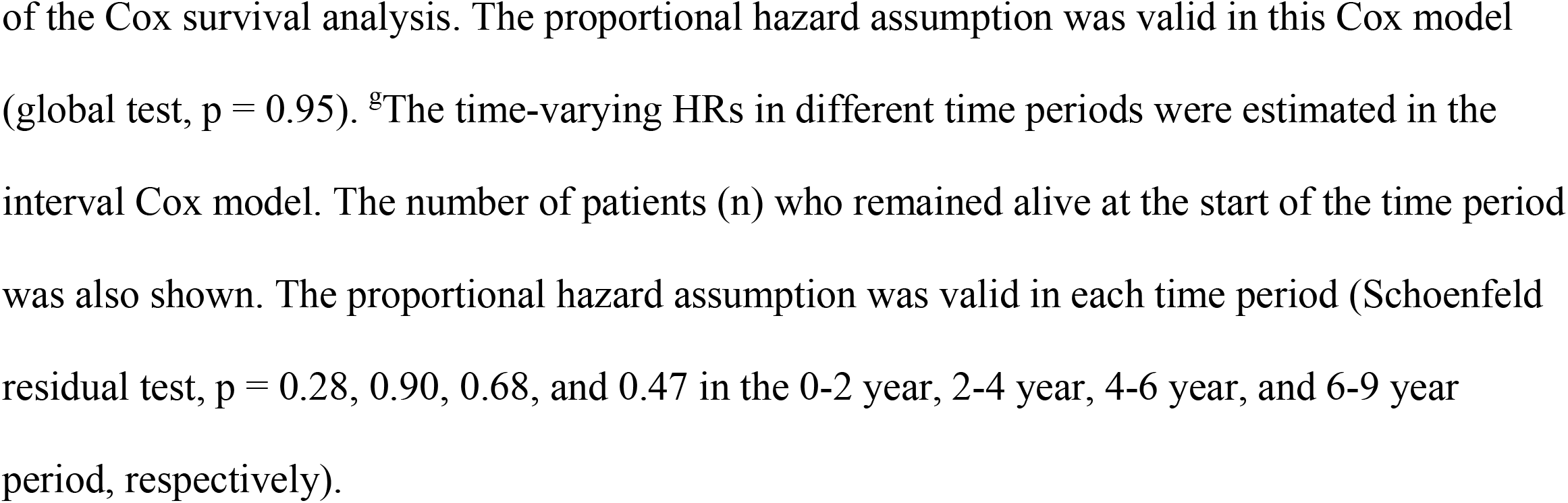
Comparison of overall mortality in the sensitivity analyses.

We performed several pre-defined subgroup analyses to test the heterogeneity among subgroups. With regard to the overall mortality, the results were consistent across most subgroups (**Figure 2**). Notably, a significant interaction between revascularization strategy and the existence of prior CAD was identified. With regard to in-hospital mortality, the results in most subgroups were largely consistent (eFigure 3 in the **Supplement**). Interestingly, possible heterogeneity of treatment effect was identified according to clinical condition within 24 hours before revascularization and prior hospitalization duration (as a proxy for frailty).

**Figure 2.**
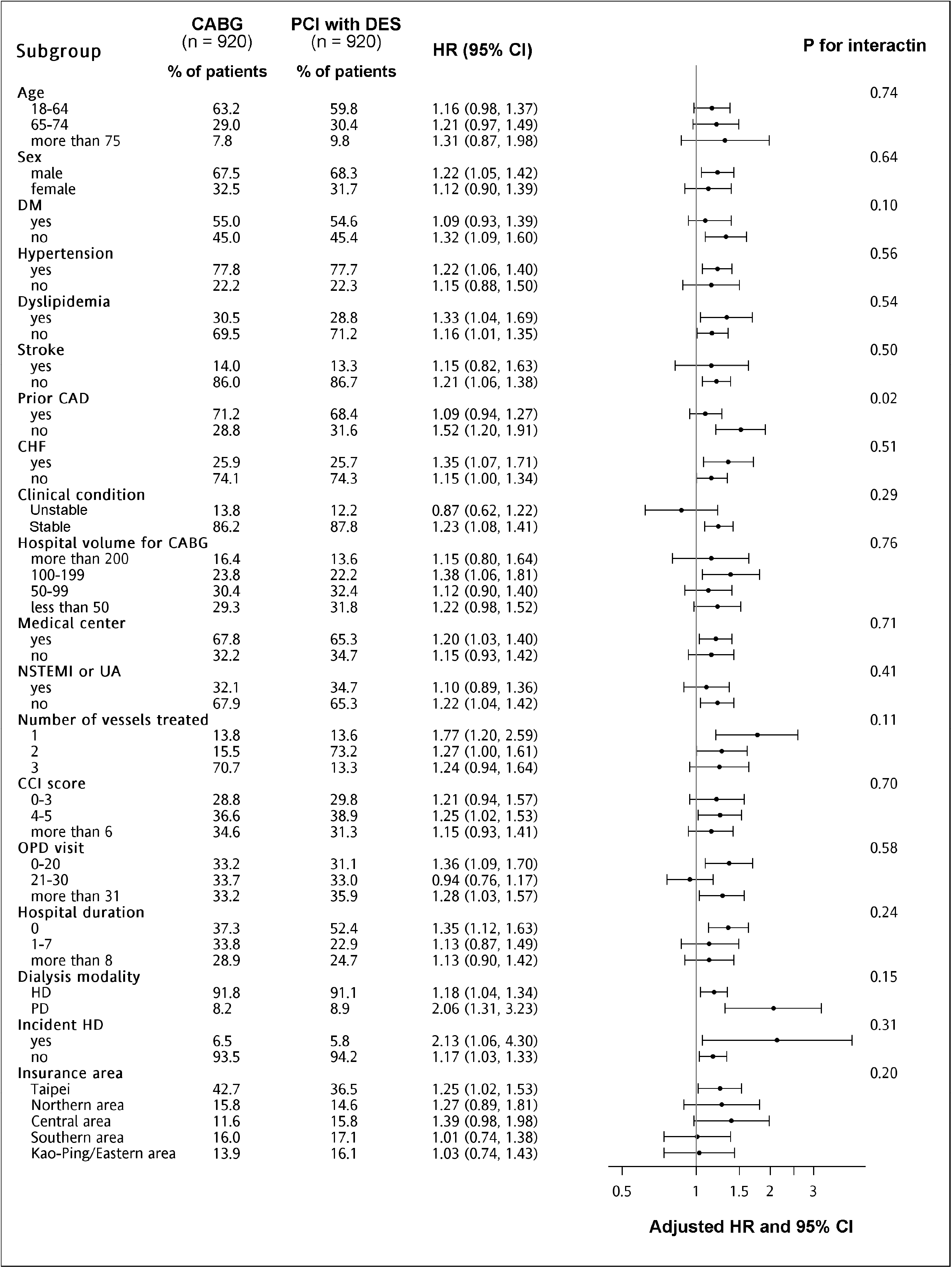
Subgroup analysis of overall survival in the matched cohort. The adjusted HRs for overall survival in the multivariable Cox proportional hazard model according to pre-specified subgroups were shown. In each subgroup, the percentage of patients from CABG (n = 920) and PCI with DES (n =920) groups were specified. Significant heterogeneity of effect was identified according to the existence of prior CAD and the frequency of prior OPD visits. Abbreviation: CAD, coronary artery disease; CCI, Charlson comorbidity index; CHF, congestive heart failure; CI, confidence interval; DM, diabetes mellitus; HD, hemodialysis; HR, hazard ratio; NSTEMI, non-ST elevation myocardial infarction; OPD, outpatient department; PD, peritoneal dialysis; UA, unstable angina

## Discussion

In our cohort of Taiwan ESKD patients, in contrast to prior observed long-term survival benefits with CABG,^9^ we found that PCI with DES is associated with lower in-hospital mortality and better long-term survival.

### The association of different revascularization strategies and in-hospital mortality

Compared with PCI with DES, patients receiving CABG had increased in-hospital mortality and increased length of hospitalization. The findings are consistent with prior observational studies.^17,23-26^ In our cohort, the crude in-hospital mortality rates in the unmatched cohort were 11.8% and 2.3% in the CABG and PCI with DES groups, respectively (eTable 1 in the **Supplement**). The duration of hospitalization was 20 (14-30) and 3 (2-8) days in the CABG and PCI with DES groups, respectively. The in-hospital mortality rate and duration of hospitalization in our study were comparable to prior reports of ESKD patients receiving CABG in the US (in-hospital mortality rate 5.4% ∼ 31%, duration of hospitalization 13 ∼ 25 days).^25^ It is likely that the increased in-hospital mortality and hospitalization duration arose from surgery-related perioperative mortality and complications in frail and heavily comorbid ESKD patients.

### The association of different revascularization strategies and long-term mortality

We found that PCI with DES was associated with better long-term survival than CABG in Taiwan ESKD patients. Even after excluding patients who died during the index hospitalization in the sensitivity analysis (**Table 3**), PCI with DES was non-inferior to CABG in long-term survival. The Interval Cox model suggested the survival benefit in the PCI with DES group may be largely derived from the reduction of early mortality after revascularization. However, PCI with DES was associated with higher risks for composite outcomes comprising death and repeat revascularization.

Notably, the cumulative 5-year survival probability of ESKD patients after revascularization was much higher in our cohort (CABG 38.7% and PCI with DES 41.9% in the matched cohort, **Figure 1**), compared with reports from URSRD (CABG 28% and PCI with DES 24%).^17^ Interestingly, in one Japanese cohort investigating mortality risk after revascularization in ESKD patients, the cumulative 5-year all-cause mortality (49.9% after CABG vs. 52.3% after PCI with DES) was very similar to our study (55.0% after CABG vs. 50.0% after PCI with DES, eTable 1 in the **Supplement**).^23^ The cause of improved long-term survival in Taiwan and Japan ESKD patients is not clear. The reason for the absence of long-term survival benefits associated with CABG is also unclear. Both biological and non-biological factors may contribute.

International comparison indicates a marked variation in overall survival in dialysis patients, which may influence the comparative effectiveness of CABG and PCI with DES.^27-29^ According to Dialysis Outcomes and Practice Patterns Study (DOPPS), the crude mortality rate from 2002 to 2008 was 18.1, 15.6, and 5.2 deaths per 100 patient-years in the US, the United Kingdom, and Japan, respectively.^28^ Taiwan had a crude mortality rate of around 11.6-11.7 deaths per 100 patient-years during the same time period.^30^ Studies reported that ethnic differences may influence the treatment effect and outcome of cardiovascular diseases.^31-33^ Although CABG is associated with better long-term survival than PCI in dialysis patients according to USRDS,^9^ it is possible that different ethnic backgrounds may modify the outcomes. Interestingly, in the aforementioned USRDS study, the authors reported in their subgroup analyses that non-white non-black race was associated with a reduced benefit of CABG.^9^ In Japan, similar to our findings, cohort study and registry analysis showed that CABG was not associated with a better 5-year all-cause mortality rate than PCI in dialysis patients.^23,26^

Notably, although PCI with DES was associated with better long-term survival in our study, repeat revascularization, subsequent ACS, and repeat hospitalization developed more frequently, probably indicating incomplete revascularization. In line with these findings, in Japanese ESKD patients, although the long-term risks for all-cause mortality were not different in PCI and CABG groups, the risk for repeat coronary revascularization was significantly higher in the PCI group.^23,26^ Interestingly, in our cohort the survival of patients after discharge from the index hospitalization in the PCI with DES group is not worse than patients in the CABG group (**Table 3**). According to statistics from the Organization for Economic Co-operation and Development (OECD) and the Ministry of Health and Welfare in Taiwan,^34,35^ the number of acute care beds per 1,000 population was 8.0, 3.4, and 2.5 for Japan, the US, and Taiwan, respectively. Since the population density (people per square kilometer of land area) in 2020 is much higher in Taiwan (673) and Japan (345) than in the US (36),^36,37^ the acute care bed density (number of acute care beds per square kilometer of land area) is also much higher in Taiwan and Japan than in the US, which may impact the accessibility to timely coronary revascularization. Because the risk of mortality associated with recurrent ACS and incompletely treated CAD may be mitigated by timely repeated revascularization, the higher acute care accessibility in Japan and Taiwan, compared with the US, may also contribute to improved survival in the PCI with DES group. It should also be noticed that CABG might be less commonly performed in Taiwan and Japan than in the US. According to USRDS,^9^ more than 50% of ESKD patients with multivessel CAD received CABG for revascularization. However, in our study, only 41.0% of ESKD patients received multivessel revascularization with CABG (**Table 1**). Similarly, in Japan’s ESKD cohort, CABG was used as a multivessel revascularization strategy in only 27-28% of dialysis patients.^23,26^ As a result, only 19.3% of CABG in Taiwan (**Table 1**), compared with 85.1% in the US, was performed at high-volume hospitals.^38^ Whether the increased utilization of PCI over CABG in Taiwan and Japan ESKD patients influenced clinical outcomes remains unclear.

### Study limitations

Several limitations should be mentioned. First, the observational nature precludes a definite conclusion. The selection biases and unmeasured confounders both may contribute to error. Second, clinical condition (Killip stage), vessel condition including distribution of vascular territories (Syntax score),^14^ and arterial graft utilization^39,40^ were important predictors of survival in patients with CAD. However, these parameters were not available in NHIRD, and we had to adjust for other available surrogate covariates such as clinical conditions before revascularization and the number of coronary arteries treated. Third, we did not analyze the impacts of BMS employed during PCI. However, studies have shown the benefits of DES over BMS in both non-CKD^41-43^ and dialysis patients.^12,13^ Fourth, we included almost exclusively the Asian population, and the results might not be able to be generalized to other ethnic groups. Finally, we did not compare medical treatment with revascularization. However, in database study, the indication bias for the comparison of conservative treatment versus intervention may be much higher than that of different revascularization strategies.

## Conclusions

In Taiwan ESKD patients, PCI with DES was associated with lower in-hospital mortality and better long-term survival, but increased repeat revascularization and subsequent ACS compared with CABG. Future population-based studies in other countries and ethnic groups are warranted to confirm the comparative effectiveness between CABG and PCI with DES in the contemporary era.

## Data Availability

The access to National Health Insurance (NHI) Research Database in Taiwan requires application and approval by Taiwan NHI Administration.

## Acknowledgment

We thank the Applied Health Research Data Integration Service from Taiwan National Health Insurance Administration for the provision of the National Health Insurance Research Database.

## Authors’ Contributions

Szu-Yu Pan and Likwang Chen had full access to all the data in the study and take responsibility for the integrity of the data and the accuracy of the data analysis.

Concept and design: Szu-Yu Pan, Ju-Yeh Yang, and Likwang Chen.

Acquisition, analysis, or interpretation of data: All authors.

Drafting of the manuscript: Szu-Yu Pan and Likwang Chen.

Critical revision of the manuscript for important intellectual content: All authors.

Statistical analysis: Ju-Yeh Yang, Nai-Chi Teng, Yun-Yi Chen, Shi-Heng Wang, and Likwang Chen.

## Disclaimer

The interpretation and conclusion in this study do not represent the views of the National Health Insurance Administration or the National Health Research Institutes.

